# Comparative Evaluation of Machine Learning and Deep Learning Models for Early Prediction of Severe Acute Pancreatitis: A Multi-Model Study Using the 2012 Revised Atlanta Classification

**DOI:** 10.64898/2026.06.20.26356146

**Authors:** Netanel Stern

## Abstract

**Background:** Acute pancreatitis (AP) is a common gastrointestinal emergency with a subset of patients progressing to severe acute pancreatitis (SAP), which carries substantial morbidity and mortality. Current clinical severity scores such as BISAP, APACHE II, Ranson, and the Modified CT Severity Index require 24-48 hours of observation before reliable assessment is possible, limiting early triage. Machine learning (ML) approaches using routine admission laboratory values may enable earlier, more accurate prediction.

**Methods:** We evaluated 11 models spanning three architectural families—classical ML (Logistic Regression, Random Forest, Gradient Boosting), feedforward deep learning (MLP, Residual MLP, Attention MLP), and recurrent deep learning (LSTM, Stacked LSTM, Bidirectional LSTM, LSTM+Attention, CNN-LSTM) —on a Chinese AP cohort of 722 patients (585 severe, 137 mild) labelled according to the 2012 Revised Atlanta Classification. Performance was assessed via 5-fold stratified cross-validation using AUC-ROC, F1 score, sensitivity, specificity, and positive predictive value (PPV), with decision thresholds optimised for maximal F1.

**Results:** Random Forest achieved the highest AUC of 0.877 (F1=0.917, sensitivity = 96.8%, PPV=87.1%), followed closely by Gradient Boosting (AUC = 0.874, F1 = 0.918). Classical ML models consistently outperformed their deep learning counterparts. CNN-LSTM was the best-performing recurrent model (AUC = 0.777) but remained inferior to all classical approaches. LSTM-family models produced AUC values of 0.684−0.777, likely reflecting the cross-sectional tabular nature of the data, which does not naturally exploit sequential modelling.

**Conclusions:** Random Forest provides robust, high-sensitivity early prediction of SAP severity using routine admission data. The findings support the clinical utility of classical ML on structured medical data and highlight the limited added value of recurrent architectures for tabular single-encounter datasets. External prospective validation is required before clinical deployment.

## 1 Introduction

Acute pancreatitis (AP) is among the most common gastrointestinal conditions requiring hospital admission, with an estimated global annual incidence of 34 per 100,000 persons and a rising trend attributed to increasing rates of gallstone disease and alcohol use [1]. Whereas the majority of AP episodes follow a mild, self-limiting course, approximately 10-20% of patients develop severe acute pancreatitis (SAP), characterised by persistent organ failure lasting more than 48 hours [2]. SAP is associated with mortality rates of 20-40% and places extraordinary demands on intensive care resources [3, 4].

Timely identification of patients destined to develop SAP is a critical clinical challenge. Early escalation to high-dependency or intensive care, aggressive fluid resuscitation, and multidisciplinary involvement can meaningfully improve outcomes [5]. However, the established severity scoring systems—Ranson criteria, APACHE II, BISAP, and the Modified CT Severity Index (MCTSI)—all share a fundamental limitation: they require clinical data accumulated over 24 to 48 hours, or depend on contrast-enhanced CT imaging that may not be appropriate in the acute setting [6, 7, 8]. There is therefore an unmet clinical need for prediction tools that can stratify severity at the time of hospital admission using readily available information.

Machine learning (ML) approaches have been applied with increasing frequency to clinical prediction tasks, including severity stratification in sepsis, acute kidney injury, and cardiovascular disease [9]. In the context of AP, several groups have reported promising results from logistic regression, ensemble tree methods, and neural network architectures applied to administrative and laboratory data [10, 11, 12]. Nevertheless, no consensus has emerged on which modelling family is best suited to this prediction task, and rigorous head-to-head comparisons of classical ML against deep learning variants—particularly recurrent architectures—remain sparse.

The present study addresses this gap with a systematic, multi-model evaluation of 11 predictors spanning three architectural families: classical ML (Logistic Regression, Random Forest, Gradient Boosting), standard feedforward deep learning (MLP, Residual MLP, Attention MLP), and recurrent deep learning (LSTM and four variants). All models were trained and evaluated on an established Chinese AP cohort labelled according to the 2012 Revised Atlanta Classification (RAC), using 5-fold stratified cross-validation [2]. Our objectives were: (i) to determine which modelling approach maximises discriminative performance for early SAP prediction; (ii) to evaluate the suitability of recurrent architectures for tabular single-encounter clinical data; and (iii) to release an open-source implementation to support reproducibility and future research.

## 2 Methods

### 2.1 Study Design and Ethical Considerations

This was a retrospective, de-identified computational study. The dataset used was assembled from a previously published Chinese AP cohort; no new patient contact or data collection was conducted. Data were fully anonymised prior to use. Accordingly, this study did not require prospective ethics approval; it was conducted under the research exemption applicable to analysis of pre-existing de-identified datasets. All analyses were performed solely for research purposes and the resulting models are not intended for clinical decision-making.

### 2.2 Dataset

The cohort comprised 722 AP patients. Severity labels were assigned according to the 2012 Revised Atlanta Classification [2], which defines SAP by the presence of persistent (>48 h) single or multi-organ failure. Of 722 patients, 585 (81.0%) met criteria for SAP and 137 (19.0%) were classified as mild or moderately severe AP. The pronounced class imbalance reflects the case-mix of tertiary referral centres in China, where severely ill patients are disproportionately represented. Class imbalance was explicitly accounted for in model training through stratified cross-validation and class-weighted loss functions where applicable.

### 2.3 Features

Input features comprised routine admission laboratory values and basic clinical measurements obtainable at or shortly after emergency department presentation, including: white blood cell count (WBC), C-reactive protein (CRP), serum creatinine, blood urea nitrogen (BUN), fasting blood glucose, serum calcium, lactate dehydrogenase (LDH), serum albumin, haematocrit, age, body mass index (BMI), heart rate, and systolic blood pressure. Features were standardised (zero mean, unit variance) prior to model training. Missing values were imputed using median imputation within each cross-validation training fold to prevent data leakage.

### 2.4 Cross-Validation Strategy

Performance was estimated using 5-fold stratified cross-validation, preserving the SAP/non-SAP ratio in each fold. All preprocessing transformations (feature scaling, imputation) were fit exclusively on training folds and applied to the corresponding held-out fold. Final performance metrics represent the mean across all five held-out test folds.

### 2.5 Models Evaluated

Eleven models were evaluated across three families:

1. **Classical ML:** Logistic Regression (L2 regularisation), Random Forest (RF; 500 estimators, balanced class weights), and Gradient Boosting (GB; 200 estimators, learning rate 0.05).
2. **Feedforward Deep Learning:** Three-layer MLP with ReLU activations and dropout; Residual MLP incorporating skip connections; Attention MLP augmented with a self-attention mechanism over feature embeddings.
3. **Recurrent Deep Learning:** Standard LSTM; Stacked (two-layer) LSTM; Bidirectional LSTM; LSTM with additive attention; CNN-LSTM (one-dimensional convolutional feature extraction followed by an LSTM layer).

Deep learning models were trained using the Adam optimiser with binary cross-entropy loss and early stopping based on validation AUC. Tabular features were reshaped to a sequence of length one for recurrent models, as the data are cross-sectional (single time-point) rather than longitudinal.

### 2.6 Threshold Optimisation and Performance Metrics

For each model, the decision threshold was selected to maximise the F1 score on the training portion of each cross-validation fold, then applied to the corresponding test fold. Performance metrics reported are: AUC-ROC (primary discriminative measure), F1 score, sensitivity (recall), specificity, and positive predictive value (PPV; precision). Given the clinical priority of minimising missed SAP diagnoses (i.e., false negatives), sensitivity was treated as a secondary primary outcome.

### 2.7 Implementation

All models were implemented in Python 3.10. Classical ML models used scikit-learn 1.3; deep learning models used TensorFlow 2.13/Keras. Source code, preprocessing pipelines, and trained model artefacts are openly available at https://github.com/netanelcyber/penuX/tree/mimic3%2B4/PenuX-AP-Severity.

## 3 Results

### 3.1 Overall Discriminative Performance

Table 1 summarises performance metrics for all 11 models. Random Forest (RF) achieved the highest AUC-ROC of 0.877, followed by Gradient Boosting (GB) at 0.874. Logistic Regression produced an AUC of 0.817. Among feedforward deep learning models, the standard MLP performed best (AUC = 0.836), followed by Residual MLP (0.804) and Attention MLP (0.784). Recurrent models showed the lowest discriminative performance overall, with AUC values ranging from 0.684 (LSTM+Attention) to 0.777 (CNN-LSTM).

**Table 1:** Performance metrics for all 11 models evaluated on the Chinese AP cohort (n = 722; 5-fold stratified cross-validation). Threshold selected to maximise F1 on the training portion of each fold. ‘—’ indicates that specificity/PPV are not reliably estimable due to threshold collapse.

| Model | Family | AUC | F1 | Sens. | Spec. | PPV | Threshold |
| --- | --- | --- | --- | --- | --- | --- | --- |
| Logistic Regression | Classical ML | 0.817 | 0.907 | 93.8% | 43.8% | 87.7% | 0.575 |
| **Random Forest** | Classical ML | **0.877** | 0.917 | 96.8% | 38.7% | 87.1% | 0.535 |
| Gradient Boosting | Classical ML | 0.874 | **0.918** | 97.1% | 38.0% | 87.0% | 0.350 |
| MLP (3-layer) | Feedforward DL | 0.836 | 0.909 | 96.9% | 24.8% | 84.6% | 0.282 |
| Residual MLP | Feedforward DL | 0.804 | 0.912 | 97.8% | 28.5% | 85.4% | 0.203 |
| Attention MLP | Feedforward DL | 0.784 | 0.909 | 98.3% | 23.4% | 84.6% | 0.418 |
| LSTM | Recurrent DL | 0.699 | 0.895 | 100% | — | — | 0.300 |
| Stacked LSTM | Recurrent DL | 0.705 | 0.898 | 98.3% | — | — | 0.489 |
| Bidirectional LSTM | Recurrent DL | 0.715 | 0.898 | 99.5% | — | — | 0.318 |
| LSTM+Attention | Recurrent DL | 0.684 | 0.896 | 100% | — | — | 0.172 |
| **CNN-LSTM** | Recurrent DL | 0.777 | 0.897 | 100% | — | — | 0.082 |

### 3.2 Sensitivity and Clinical Relevance

Given the potential consequences of missing a SAP diagnosis, sensitivity was a key performance consideration. At its optimised threshold of 0.535, RF correctly identified 96.8% of SAP cases (estimated 566 true positives, 19 false negatives in the cohort). GB achieved the highest F1 of 0.918 with sensitivity of 97.1%. Attention MLP achieved the highest sensitivity among deep learning models (98.3%), while all CNN-LSTM and several LSTM variants reached 100% sensitivity, albeit with very low decision thresholds (e.g., CNN-LSTM threshold = 0.082) and correspondingly poor specificity metrics not computable in a meaningful clinical sense.

### 3.3 Classical ML versus Deep Learning

Classical ML models consistently outperformed deep learning counterparts on the primary discriminative metric (AUC). The RF advantage over the best recurrent model (CNN-LSTM) was 0.877 vs 0.777—a clinically meaningful difference of 0.100 AUC points. GB similarly outperformed all LSTM variants. Within the feedforward deep learning category, the standard MLP (AUC = 0.836) was competitive with classical ML, suggesting that recurrent rather than deep architectures per se is the driver of underperformance.

### 3.4 LSTM Family Performance

LSTM-family AUC values (0.684-0.777) were notably lower than classical ML and feed-forward DL. The CNN-LSTM, which applies convolutional feature extraction before the recurrent layer, partially recovered performance (AUC = 0.777) relative to vanilla LSTM (0.699). All LSTM variants achieved very high sensitivity (98.3-100%) at the cost of near-zero specificity, consistent with models defaulting toward the majority (SAP) class.

### 3.5 Summary Table

## 4 Discussion

### 4.1 Random Forest Robustness on Tabular Medical Data

The superior performance of Random Forest and Gradient Boosting in this study is consistent with a growing body of evidence that ensemble tree methods outperform deep learning on structured (tabular) biomedical data, particularly in moderate-sized cohorts [13, 14]. Random forests are robust to feature scaling, naturally handle feature interactions, and provide implicit feature selection through bagging and random subspace sampling [15]. In clinical datasets of the size evaluated here (n = 722), the inductive bias of tree ensembles—which does not assume smoothness or sequential dependency in the feature space—is well matched to laboratory data that are independently distributed across samples.

### 4.2 Clinical Priority of Sensitivity

In clinical triage, the asymmetry of error consequences is paramount. A missed SAP diagnosis (false negative) results in inadequate monitoring, delayed resuscitation, and potentially preventable mortality, while a false positive results primarily in unnecessary ICU-level resource utilisation. The RF at threshold 0.535 achieves 96.8% sensitivity, capturing the large majority of true SAP cases while maintaining a PPV of 87.1%. This compares favourably with published BISAP and APACHE II sensitivity values of approximately 60-80% for SAP within 24 hours of admission [6, 7]. However, direct comparison is limited by the absence of BISAP/APACHE II validation on this same cohort, a noted limitation of the present work.

### 4.3 LSTM Underperformance: Tabular Data and Architectural Mismatch

The markedly lower AUC of LSTM-family models (0.684-0.777) relative to classical ML and feedforward DL warrants careful interpretation. Recurrent architectures were developed to exploit sequential temporal structure in time-series data such as electronic health record trajectories, waveforms, or longitudinal laboratory measurements [9, 16]. When applied to cross-sectional tabular data—where each patient is represented by a single feature vector with no inherent ordering or sequential dependency—recurrent models must be artificially reshaped (e.g., treating features as a one-step sequence), eliminating the architectural benefit for which they were designed. The CNN-LSTM partially mitigated this by using convolutional layers to extract local feature interactions before the recurrent stage, explaining its marginal advantage over vanilla LSTM. The consistent 100% sensitivity of several LSTM models, combined with threshold collapse to near-zero values, indicates these models are essentially predicting the majority class (SAP) for all inputs—a degenerate solution that inflates sensitivity while providing no real discriminative utility.

### 4.4 Class Imbalance

The 81:19 SAP-to-non-SAP ratio in this cohort reflects referral bias inherent to tertiary pancreatitis centres in China. This imbalance may inflate absolute sensitivity metrics across all model types and limits generalisability to community or mixed-acuity settings where mild AP predominates [1]. Future work should evaluate model calibration and net reclassification improvement in more balanced, community-based cohorts.

### 4.5 Limitations

Several limitations must be acknowledged. First, all analysis was conducted on a single Chinese cohort; ethnic, dietary, and aetiological differences in AP epidemiology mean that performance may not generalise to Western or mixed-aetiology populations. Second, no external validation set was available; 5-fold cross-validation, while standard, may overestimate generalisation performance compared with true hold-out external validation. Third, the class imbalance (81% severe) may have driven models toward high-sensitivity, low-specificity operating points. Fourth, we did not include a direct comparison with clinical scoring systems (BISAP, APACHE II, Ranson) on this cohort, limiting the ability to contextualise clinical utility. Fifth, feature importance analysis was not reported here but is a planned extension. Finally, the retrospective design precludes causal inference.

### 4.6 Future Directions

Priorities for future work include: prospective validation in multi-centre cohorts with balanced severity distributions; head-to-head comparison with established clinical scoring systems on the same patient sample; systematic feature importance analysis to identify the most predictive admission biomarkers; exploration of calibrated probability outputs for clinical risk communication; and investigation of whether longitudinal (multi-time-point) data collection would rehabilitate recurrent architectures in this setting.

## 5 Conclusion

In this comparative evaluation of 11 machine learning and deep learning models for early severity prediction in acute pancreatitis, Random Forest achieved the best overall discriminative performance (AUC = 0.877) using routine admission laboratory values, with Gradient Boosting performing comparably (AUC = 0.874). Classical ensemble methods consistently outperformed feedforward and recurrent deep learning architectures. LSTM-family models were poorly suited to this cross-sectional tabular setting, achieving AUC values of 0.684-0.777 and exhibiting threshold collapse toward universal positive prediction. The high sensitivity of RF (96.8% at threshold 0.535) is clinically meaningful for early identification of patients requiring escalated care. An open-source implementation is freely available at https://github.com/netanelcyber/penuX/tree/mimic3%2B4/PenuX-AP-Severity to facilitate reproducibility and adaptation. Prospective, multi-centre external validation is required before any clinical deployment.

### Disclaimer

This study is for research purposes only and is not intended for clinical use. The predictive models described have not undergone prospective clinical validation and must not be used to guide individual patient management.

## Data Availability

All data produced are available online at: https://github.com/longshike/LNN-for-SAP-Prediction

https://github.com/longshike/LNN-for-SAP-Prediction

## Data Availability

The de-identified dataset is not publicly distributed due to patient privacy constraints. The full analysis pipeline, model architectures, and preprocessing code are available at https://github.com/netanelcyber/penuX/tree/mimic3%2B4/PenuX-AP-Severity.

## Code Availability

Open-source Python implementation: https://github.com/netanelcyber/penuX/tree/mimic3%2B4/PenuX-AP-Severity. Requires Python 3.10, scikit-learn 1.3, TensorFlow 2.13, and standard scientific Python libraries (NumPy, pandas, Matplotlib).

## Author Contributions

N.S.: Conceptualisation, data curation, model development, analysis, manuscript writing.

## Acknowledgements

The author thanks the open-source Python and ML communities whose tools made this work possible.

## References

[1] Lankisch PG, Apte M, Banks PA. Acute pancreatitis. Lancet. 2015;386(9988):85–96. 10.1016/S0140-6736(14)60649-8

[2] Banks PA, Bollen TL, Dervenis C, et al.; Acute Pancreatitis Classification Working Group. Classification of acute pancreatitis—2012: revision of the Atlanta classification and definitions by international consensus. Gut. 2013;62(1):102–111. 10.1136/gutjnl-2012-302779

[3] Petrov MS, Shanbhag S, Chakraborty M, Phillips ARJ, Windsor JA. Organ failure and infection of pancreatic necrosis as determinants of mortality in patients with acute pancreatitis. Gastroenterology. 2010;139(3):813–820. 10.1053/j.gastro.2010.06.010

[4] Garg PK, Singh VP. Organ failure due to systemic injury in acute pancreatitis. Gastroenterology. 2019;156(7):2008–2023. 10.1053/j.gastro.2018.12.041

[5] Tenner S, Baillie J, DeWitt J, Vege SS; American College of Gastroenterology. American College of Gastroenterology guideline: management of acute pancreatitis. Am J Gastroenterol. 2013;108(9):1400–1415. 10.1038/ajg.2013.218

[6] Papachristou GI, Muddana V, Yadav D, et al. Comparison of BISAP, Ranson’s, APACHE-II, and CTSI scores in predicting organ failure, complications, and mortality in acute pancreatitis. Am I Gastroenterol. 2010;105(2):435–441. 10.1038/ajg.2009.622

[7] Wu BU, Johannes RS, Sun X, Tabak Y, Conwell DL, Banks PA. The early prediction of mortality in acute pancreatitis: a large population-based study. Gut. 2008;57(12):1698–1703. 10.1136/gut.2008.152702

[8] Bollen TL, Singh VK, Maurer R, et al. A comparative evaluation of radiologic and clinical scoring systems in the early prediction of severity in acute pancreatitis. Am J Gastroenterol. 2012;107(4):612–619. 10.1038/ajg.2011.438

[9] Rajpurkar P, Chen E, Banerjee O, Topol EJ. AI in health and medicine. Nat Med. 2022;28(1):31–38. 10.1038/s41591-021-01614-0

[10] Qiu Q, Nian YJ, Guo Y, et al. Development and validation of three machine-learning models for predicting multiple organ failure in moderately severe and severe acute pancreatitis. BMC Gastroenterol. 2021;21(1):367. 10.1186/s12876-021-01946-6

[11] Mofidi R, Duff MD, Madhavan KK, Garden OJ, Parks RW. Identification of severe acute pancreatitis using an artificial neural network. Surgery. 2007;141(1):59–66. 10.1016/j.surg.2006.07.019

[12] Mounzer R, Langmead CJ, Wu BU, et al. Comparison of existing clinical scoring systems to predict persistent organ failure in patients with acute pancreatitis. Gastroenterology. 2012;142(7):1476–1482. 10.1053/j.gastro.2012.03.005

[13] Grinsztajn L, Oyallon E, Varoquaux G. Why tree-based models still outperform deep learning on tabular data. Adv Neural Inf Process Syst. 2022;35:507–520. arXiv:2207.08815

[14] Shwartz-Ziv R, Armon A. Tabular data: deep learning is not all you need. Inf Fusion. 2022;81:84–90. 10.1016/j.inffus.2021.11.011

[15] Breiman L. Random forests. Mach Learn. 2001;45(1):5–32. 10.1023/A:1010933404324

[16] Shickel B, Tighe PJ, Bihorac A, Rashidi P. Deep EHR: a survey of recent advances in deep learning techniques for electronic health record (EHR) analysis. IEEE J Biomed Health Inform. 2018;22(5):1589–1604. 10.1109/JBHI.2017.2767063

[17] Ranson JH, Rifkind KM, Roses DF, Fink SD, Eng K, Spencer FC. Prognostic signs and the role of operative management in acute pancreatitis. Surg Gynecol Obstet. 1974;139(1):69–81.

[18] Knaus WA, Draper EA, Wagner DP, Zimmerman JE. APACHE II: a severity of disease classification system. Crit Care Med. 1985;13(10):818–829. 10.1097/00003246-198510000-00009

[19] Singh VK, Wu BU, Bollen TL, et al. A prospective evaluation of the bedside index for severity in acute pancreatitis score in assessing mortality and intermediate markers of severity in acute pancreatitis. Am J Gastroenterol. 2009;104(4):966–971. 10.1038/ajg.2009.28

[20] Chen JH, Zheng ZH, Chen XD, Chen QL, Li G. A machine learning-based severity prediction tool for the Bedside Index for Severity in Acute Pancreatitis. Front Med (Lausanne). 2021;8:652926. 10.3389/fmed.2021.652926

